# Impact of Losartan on Portal hypertension and Liver Cirrhosis: A Systematic Review

**DOI:** 10.1101/2024.09.25.24314392

**Authors:** Deepti Avasthi, Salil Avasthi

## Abstract

**Background:** Portal hypertension, a complication of chronic liver disease, results from elevated pressure between the portal vein and the inferior vena cava. While non-selective beta-blockers are established for reducing portal pressure, the efficacy of losartan, an angiotensin II receptor blocker, remains debated. This study evaluated losartan’s impact on portal pressure and clinical outcomes in patients with cirrhosis and portal hypertension.

**Objectives:** To appraise evidence on the role of losartan in reducing portal pressure and associated clinical outcomes in cirrhotic patients with portal hypertension.

**Methods:** A comprehensive literature search was conducted in PubMed, Cochrane Library, Medline, and Web of Science. All the research and review of literature was conducted within the time period from August 20th, 2024 till August 31st, 2024 (within 1 month of submission of the paper). The Risk of Bias Visualization Tool (Robvis 2.0) and ROBINS-I were used to assess study quality. Data was extracted and analyzed using Microsoft Excel.

**Results:** Among 426 potential studies, 12 met the inclusion criteria. Both losartan and propranolol reduced hepatic venous pressure gradient (HVPG), with some studies suggesting a more pronounced effect of losartan. Meta-analysis found no significant difference in HVPG reduction (p = 0.22), but losartan significantly reduced wedged hepatic venous pressure (WHVP) compared to propranolol (p = 0.03). Losartan also affected mean arterial pressure, renal function, and hepatic fibrosis.

**Conclusions:** Losartan shows potential in treating portal hypertension by reducing portal pressure and fibrosis. It may be particularly beneficial for patients unresponsive to beta-blockers, addressing both hemodynamic and structural components, and improving sodium handling in complex cases.

## Introduction

Portal hypertension is a serious complication of chronic liver diseases that can lead to life-threatening conditions like variceal bleeding and ascites. It arises from an increased pressure gradient between the portal vein and the inferior vena cava, often due to elevated intrahepatic vascular resistance and splanchnic vasodilation (1). This condition can cause liver distortions, secondary fibrosis, and elevated portal blood pressure, particularly in cirrhosis (2). The pathogenesis of liver cirrhosis is strongly associated with cardiovascular morbidity and shares a common pathologic pathway (3).

Current treatment primarily involves non-selective beta-blockers to reduce portal pressure (4). However, the renin-angiotensin system presents another therapeutic target, with Angiotensin II receptor blockers like losartan showing promise in experimental studies (5,6). Research has investigated losartan’s effects on portal pressure and clinical outcomes in cirrhotic patients, revealing mixed results regarding its impact on hepatic venous pressure gradient (HVPG), likely due to variations in patient demographics and study designs.

Angiotensin II receptor blocker significantly influences the pathogenesis of hepatic fibrosis, vasoconstriction within the liver sinusoids and sodium balance in portal hypertension (7). However, angiotensin receptor blockers can cause complications such as hypotension and renal impairment (8). The pathogenesis of fibrosis could be reduced by losartan because it could potentially inhibit the angiotensin II receptors and offer a new treatment opportunity for liver cirrhosis.

This study aims to evaluate losartan’s effect on HVPG, systemic hemodynamics, renal function, and clinical events like variceal bleeding and ascites, while also assessing the management of risks associated with its use.

### Research Objectives

This study aimed to investigate the effect of losartan on portal hypertension through a systematic review and meta-analysis of existing evidence.

### Research Questions

- How does losartan influence the progression of liver fibrosis and associated complications in patients with portal hypertension?
- What is the efficacy of losartan in reducing portal pressure in these patients?

### Methodology

The study followed the Preferred Reporting Items for Systematic Reviews and Meta-Analysis (PRISMA) guidelines (9).

### Identification and Selection of Studies

Literature was sourced from PubMed, Cochrane Library, Medline, and Web of Science focusing on losartan’s effect on portal hypertension. *(Figure 1 and supplementary table 1 and 2)*

**Figure 1:**
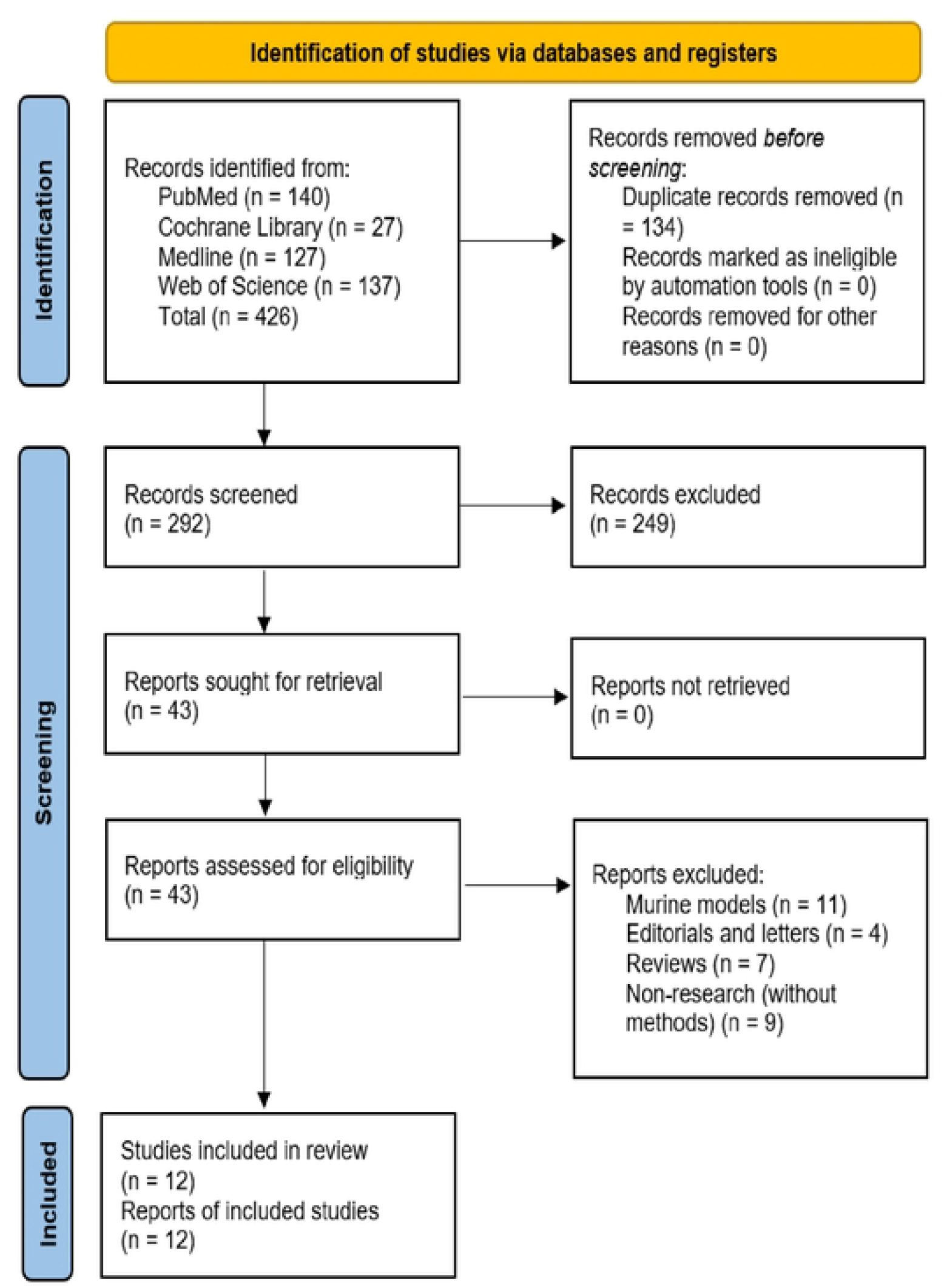
PRISMA flow diagram

### Search Strategy

Keywords used included: losartan, angiotensin II receptor blocker (ARB), portal hypertension, liver cirrhosis, and hepatic fibrosis. (*supplementary tables 1 and 2)*.

### Study Selection

Retrieved results were managed using Zotero (version 6.0.36), which helped exclude retracted records and merge duplicates.

*Figures 2, 3, 4, and 5 show the risk of bias visualization tool (Robvis 2*.*0) and the Risk of Bias in Non-randomized studies with the intervention assessment results (ROBINS)*

**Figure 2.**
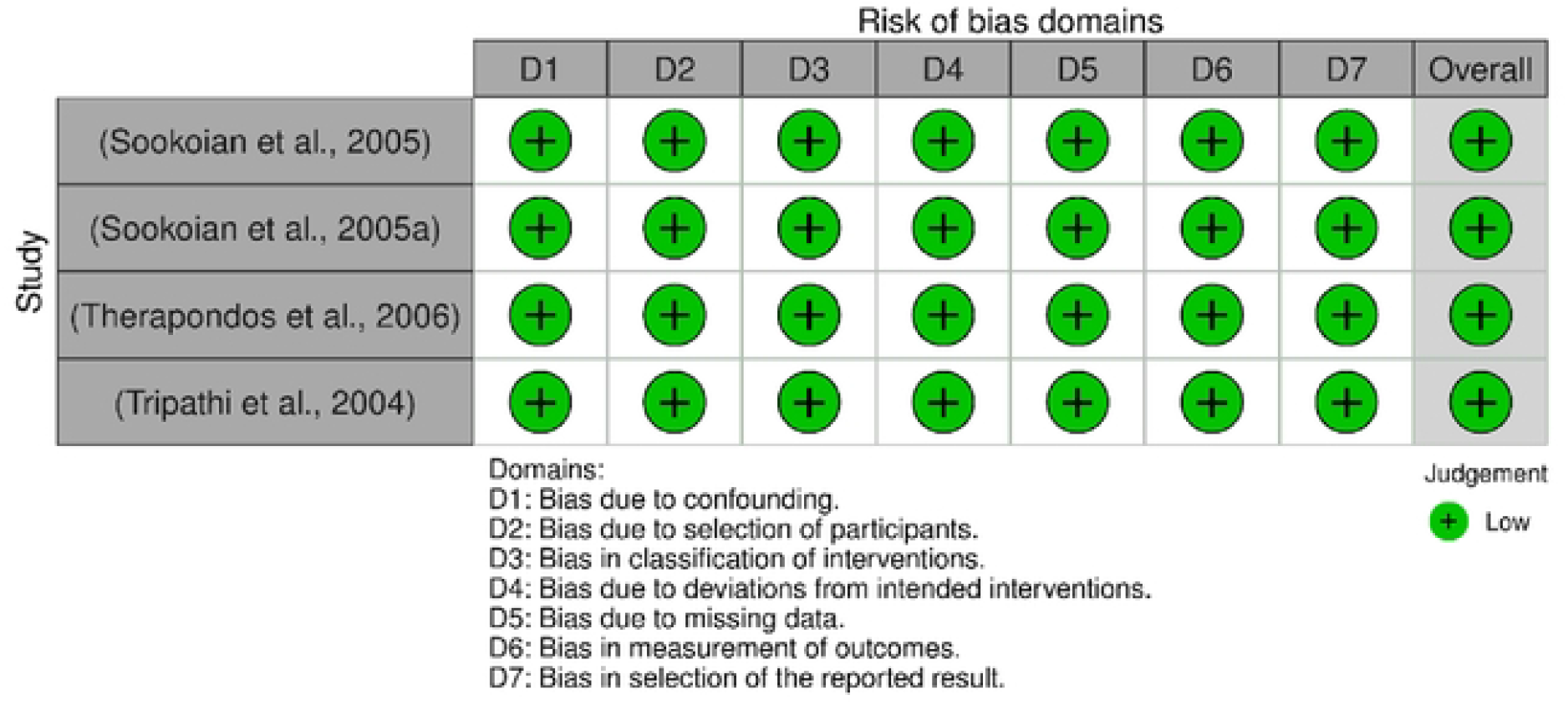
Traffic lights plot of the Risk of Bias in Non-randomized studies with the intervention assessment results (13, 18, 21, 22)

**Figure 3.**
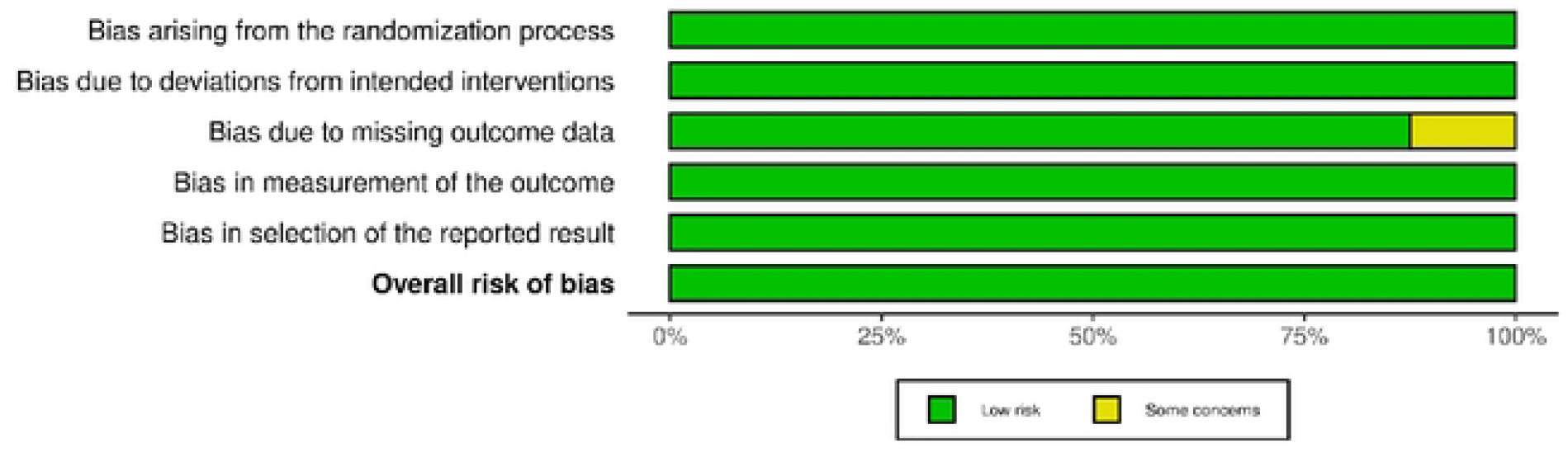
Summary plot of the Risk of Bias in Non-randomized studies with the intervention assessment results (13, 18, 21, 22)

**Figure 4.**
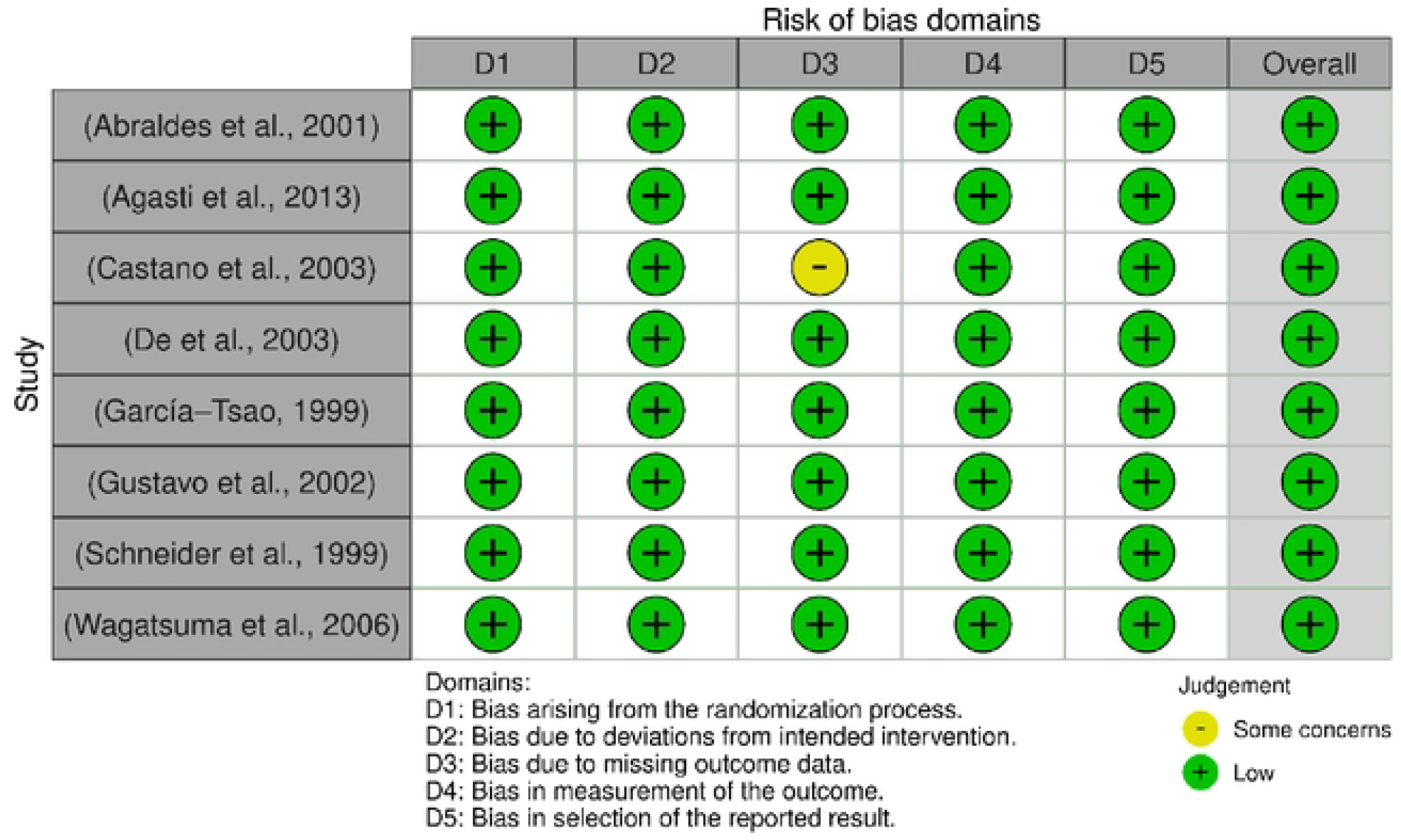
Traffic lights plot of the risk of bias visualization tool assessment results (14, 15, 16, 17, 19, 20, 23, 31)

**Figure 5.**
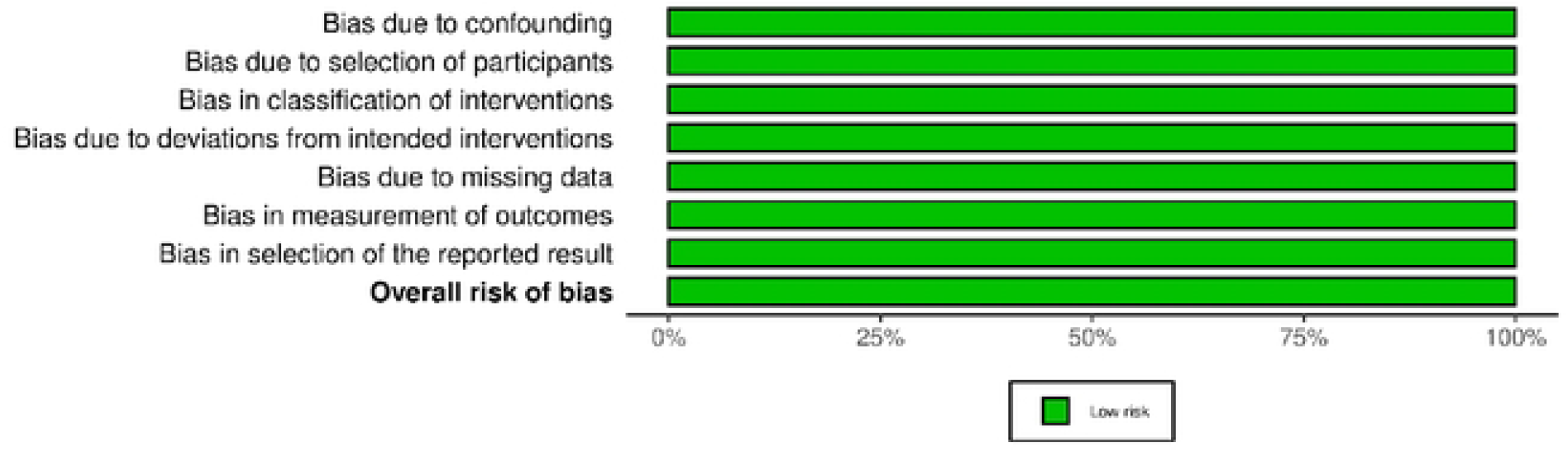
Summary plot of the risk of bias visualization tool assessment results (14, 15, 16, 17, 19, 20, 23, 31)

### Eligibility Criteria

Research included in the study adhered to modified PICOS criteria (10):

- Population (P): Patients with portal hypertension.
- Intervention (I): Losartan.
- Comparison (C): None defined.
- Primary Outcomes (O): Portal pressure, response rates, disease progression.

#### Study Design (S)

Quantitative, qualitative, and mixed methods.

### Inclusion Criteria

Peer-reviewed original research articles published in English (or translatable) on losartan’s effect on portal hypertension.

### Exclusion Criteria

Excluded were non-research articles, study protocols, reviews, meta-analyses, opinion pieces, conference abstracts, and editorials.

### Methodological Quality Assessment: The risk of bias

The risk of bias was evaluated using the Robvis 2.0 tool and the Risk of Bias in Non-randomized Studies (ROBINS) (11).

All the research and review of literature was conducted within the time period from August 20th, 2024 till August 31st, 2024.

*Figures 2, 3, 4, and 5 show the risk of bias visualization tool (Robvis 2*.*0) and the Risk of Bias in Non-randomized studies with the intervention assessment results*.

### Data Extraction

Data were systematically extracted by one reviewer independently and manually into an Excel sheet using Microsoft Excel 2019, including study ID, design, settings, sample characteristics, mean age or age range, sample size, intervention details, purpose, outcome measures, and findings.

### Data Analysis

The extracted qualitative data were analyzed and reported according to the predominant themes. On the other hand, quantitative data were analyzed using Review Manager 5.4.1. An intervention review starting from the full review stage was used (12). In addition, continuous data types were used, applying inverse variance statistical, random effects analysis model, and mean difference effect measure.

### Study Selection

The literature search yielded 431 records, with 134 duplicates removed. Following the title and abstract screening, 255 records were excluded, leaving 42 articles for retrieval. Ultimately, 12 studies met eligibility criteria, as illustrated in *Figure 1*.

### Thematic Analysis of Outcomes and study characteristics: *(supplementary Table 3)*

### Changes in Hepatic Venous Pressure Gradient (HVPG) and Wedged Hepatic Venous Pressure (WHVP)

Research indicated varying effects of losartan on hepatic venous pressure gradient (HVPG), WHVP and MAP. The study conducted by (Tripathy et al). showed no significant change in HVPG after four weeks of losartan treatment, with a reduction from 15.4 ± 1.5 to 13.6 ± 1.6 mmHg (p = 0.1). There was a significant reduction in WHVP, falling from 20.3 ± 1.8 to 17.3 ± 1.8 mmHg (p < 0.05) and a significant decrease in MAP, from 97 ± 3.0 mmHg to 89 ± 4.0 mmHg (p = 0.02) (13). Another study found a significant reduction in HVPG in patients with severe portal hypertension, dropping from 24.8 ± 3.6 to 13.1 ± 4.1 mmHg (P < 0.001), compared to the control group with a minor decline from 23.9 ± 4.1 to 23.1 ± 4.2 mmHg and WHVP reduction from 22.1 ± 2.6 to 14.1 ± 2.9 mmHg (P < 0.001) in patients with moderate hypertension. The control group demonstrated a smaller reduction from 22.0 ± 2.2 to 21.4 ± 2.6 mmHg (14)

In other studies losartan reduced HVPG in moderate cases by 46.8%, from 17.9 ± 1.4 to 10.0 ± 2.7 mmHg (15). Comparatively, losartan reduced HVPG from 15.6 ± 4.2 mmHg to 11.8 ± 3.5 mmHg, while propranolol reduced it from 16.4 ± 4.1 to 13.1 ± 3.6 mmHg, showing a more significant reduction of -10% ± 11% (P = 0.003) compared to losartan’s non-significant change of -2% ± 12% (16).The study conducted by De Buke et al. showed losartan decreased HVPG from 19.21 ± 3.82 to 14.15 ± 4.91 mmHg, while propranolol reduced it from 18.7 ± 3.77 to 15.45 ± 5.35 mmHg. Losartan also reduced WHVP from 32.42 ± 6.61 to 28.31 ± 5.09 mmHg, and propranolol lowered it from 34.55 ± 5.41 to 32.75 ± 8.13 mmHg (17). Losartan significantly decreased MAP from 97 ± 3.0 to 89 ± 4.0 mmHg (p = 0.02) and from 90.9 ± 5.5 to 87.4 ± 4.6 mmHg in 2 other studies who compared its effect with propanol. A reduction in systolic arterial pressure from 134 ± 22.7 to 124 ± 18.1 mmHg was noted, along with minor increases in diastolic pressure and MAP. In comparison, propranolol led to significantly lower HVPG and mean arterial blood pressure (MABP), with HVPG remaining higher in the losartan group. Losartan reduced the heart rate from 79.6 ± 1.5 to 78.1 ± 2.5 BPM, while propranolol led to a larger decrease from 80.8 ± 6.2 to 68.3 ± 5.5 BPM (18,19).

### Patient Demographics and Genetic Factors

The study conducted by (Sookoian et al) on genetic polymorphism provided some insights on the the variable effect of losartan in different genotypes. According to this study losartan significantly decreased HVPG in portal hypertension patients with genotype AA from 15.7 ± 4.3 to 10.6 ± 3.4, in contrast to patients with genotypes AC and CC, who showed minor reductions from 15.9 ± 1.6 to 15.4 ± 2.8. A similar trend was observed in WHVP, where genotype AA patients showed significant improvement from 26.0 ± 4.4 to 21.9 ± 6.1 mmHg, while no significant changes were noted in patients with AC and CC genotypes.

### Impact on Renal Markers and sodium handling

The renal function markers like glomerular filtration rate (GFR), blood urea nitrogen, and creatinine were not affected by propranolol. However, losartan significantly decreased GFR in Child B patients, particularly those with reduced MAP and systemic vascular resistance. Additionally, there were no significant changes in GFR during diuretic use (20). The effect of losartan on sodium handling in kidneys was also noted by (Tripathy et al) during their study on HVPG (quoted above). According to their findings there was no significant change in creatinine clearance after four weeks of losartan, it was recorded that losartan caused reduction of sodium excretion from 154 ± 61 mmol/day to 122 ± 36 mmol/day. Similar findings were recorded by another study losartan administration resulted in decreased proximal tubular reabsorption of sodium in both supine and erect positions, with significant decreases noted in both positions (supine: from 69.7 ± 3.1% to 63.9 ± 3.9 %, p = 0.01; erect: from 81.1 ± 1.8% to 73.8 ± 2.4%, p = 0.01) (21).

### Antifibrotic Effects

Losartan led to a decrease in the fibrosis stage, with the treated group showing a reduction of 0.64 ± 1.3 compared to an increase of 0.89 ± 1.27 in the control group. The reduction in fibrosis was further evidenced by improvements in 7 out of 14 treated patients, compared to only one out of nine in the control group. Sub-endothelial fibrosis in lobular areas significantly decreased from a baseline of 2.48 ± 1.04 to 1.00 ± 0.53 after losartan treatment, while no significant changes were observed in the control group (22).

### Management of Portal Hypertension-related Complications

According to a study conducted by Wagatsuma et al. portal hypertensive gastropathy improved in nine out of 16 patients, with an efficacy rate of 56% after losartan administration. Higher efficacy was observed with a 50mg dose of losartan (83%) compared to a 25mg dose (40%) (23).

### Safety and Adverse Effects

Losartan and propranolol were well-tolerated by over 90% of patients, though some studies reported adverse events. Transient hypotension was commonly seen after the first dose of losartan, with no recurrence during continued treatment. Mild orthostatic hypotension, gastrointestinal bleeding, and encephalopathy were also noted, with some patients developing these conditions after using either losartan or propranolol. Severe side effects included rebleeding associated with losartan, though none of the adverse events were fatal, and treatment withdrawal was necessary in only one case. In patients with severe portal hypertension, losartan also caused nausea and dizziness. The findings from the study from *Abraldes JG et al*. show that the overall incidence of adverse events was similar for losartan (28%) and propranolol (27%)

### Meta-Analysis Findings

The meta-analysis included five studies (study numbers 14, 16, 17, 19, 20) that reported quantitative data on HVPG changes. The analysis indicated no statistically significant difference in HVPG change between the losartan treatment group and the control group (p = 0.22). High heterogeneity was noted among the included studies, with an I2 of 84%, likely due to variations in sample sizes and study designs. (Figure *6* and Figure 7)

**Figure 6:**
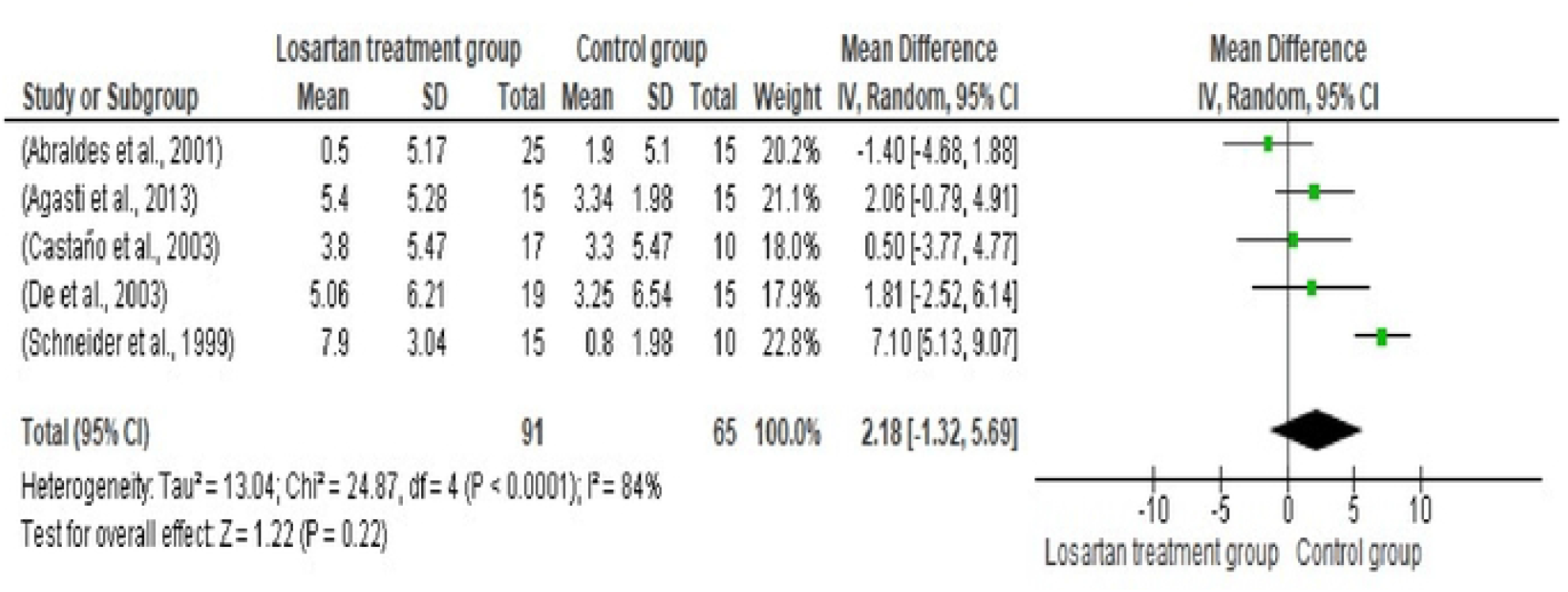
Forest plot of Hepatic Venous Pressure gradient comparison between the Losartan treatment group and the control group

**figure 7:**
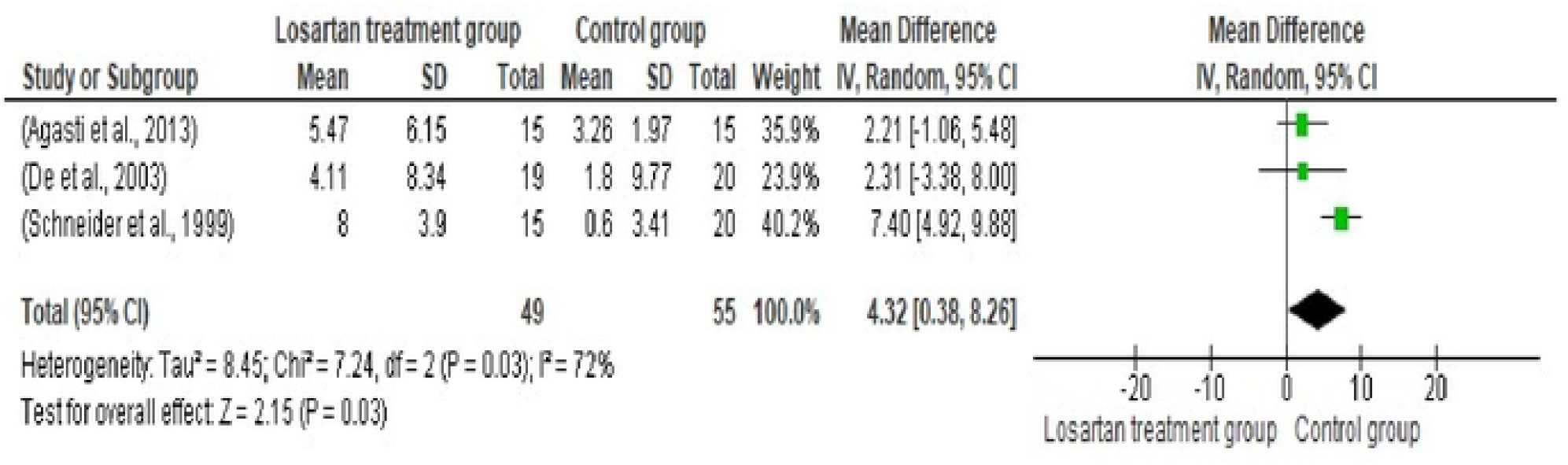
Forest plot of Wedged Hepatic Venous Pressure (WHVP) comparison between the Losartan treatment group and the control group (17, 19, 22)

Overall, this structured analysis underscores losartan’s potential efficacy in managing portal hypertension, with specific emphasis on its effects on HVPG, WHVP, and MAP, as well as its comparative efficacy against propranolol. The findings also highlight the importance of patient demographics, particularly genetic factors, and renal function in treatment outcomes.

## Discussion

This study assessed the effects of losartan on portal hypertension. The findings of this systematic review and meta-analysis indicate that managing the condition is challenging. The variable results in terms of the effectiveness of losartan for lowering HVPG across studies relate directly to the variability in the severity and baseline characteristics of portal hypertension. Some studies showed marked reductions in HVPG, while others showed negligible changes, resulting in no statistically significant overall change in HVPG in the meta-analysis. These differences could be attributed to factors such as the variation in liver disease type and extent, initial portal hypertension severity, and differences in treatment duration and dosages (24).

Further investigation into losartan’s hemodynamic effects revealed changes in WHVP. However, the impact was not statistically significant, highlighting that portal hypertension’s pathophysiology is multifactorial, with various mechanisms influencing the initial improvements. Most studies noted decreases in mean arterial pressure, a potential concern for cirrhotic patients with fragile hemodynamic conditions. Thus, losartan therapy must be carefully adjusted to avoid systemic hypotension.

Losartan also showed therapeutic potential in mitigating liver disease-related fibrosis, aligning with preclinical findings that suggest losartan could inhibit hepatic stellate cell activation (25). This variability in effect corresponds with previous studies on the Renin-Angiotensin System in portal hypertension, linking it to complex pathways, high intrahepatic pressure, and splanchnic vasodilation (26).

Genetic factors, such as the AT1R A1166C polymorphism, may influence losartan effectiveness, suggesting personalized treatment options for portal hypertension (27). The significant plasticity of hepatic circulation leads to compensatory reactions that might minimize losartan’s initial hemodynamic effects (28), which does not guarantee long-term outcomes.

Dose optimization remains inconclusive, as higher doses reduce portal pressure but increase the risk of systemic hypotension. The timing of intervention is also crucial, with early intervention potentially offering benefits before disease progression.

The safety profile of losartan showed good tolerability, though episodes of hypotension, reduced renal function, and gastrointestinal hemorrhage were reported. These findings correlate with the Renin-Angiotensin System inhibition effects in cirrhotic patients emphasizing careful patient evaluation and monitoring (29,30).

Overall, the variability in response underscores the need to consider multiple factors in managing portal hypertension, including patient characteristics, disease etiology and severity, and genetic predispositions. Losartan’s antifibrotic effects point to potential therapeutic approaches in the disease’s early stages. Combination therapy targeting hemodynamic and fibrotic features might enhance treatment outcomes. However, safety concerns necessitate meticulous patient selection, especially for those with unstable hemodynamics. Further studies should explore the factors affecting losartan response, optimal dosing, and intervention timing in liver disorders.

### Limitations of the study

The heterogeneity among included studies regarding patient populations and dosing impacts the generalizability of the findings. In addition, the variability in disease etiology, severity, and baseline portal pressures across studies limit direct comparisons through meta-analyses.

Moreover, the lack of data on specific subgroups, such as patients with different stages of liver disease or various comorbidities, limits the generalizability of the findings of losartan’s efficacy to all subgroups.

## Conclusion

This study highlights losartan’s potential efficacy in treating portal hypertension, particularly for non-responders to conventional beta-blockers. Its antifibrotic properties and ability to reduce portal pressure highlight its potential to address both hemodynamic and structural aspects of the disease, especially in the early stage of liver cirrhosis.

Moreover, given the high incidence of cardiomyopathy in cirrhotic patients, losartan’s effects on mean arterial pressure and natriuresis could also be relevant in managing associated cardiac comorbidities. Further research is needed to fully explore losartan’s comprehensive therapeutic benefits in this complex patient population, particularly focusing on its dual role in liver and cardiovascular health.

## Data Availability

All relevant data are within the manuscript and its Supporting Information files.

na

## Notes

### Competing Interest Statement

The authors have declared that no competing interests exist.

### Clinical Trial

na

### Clinical Protocols

naga

### Funding Statement

The author(s) received no specific funding for this work.

